# Detection and stability of SARS-CoV-2 fragments in wastewater: Impact of storage temperature

**DOI:** 10.1101/2021.02.22.21250768

**Authors:** Rudolf Markt, Markus Mayr, Evelyn Peer, Andreas O. Wagner, Nina Lackner, Heribert Insam

**Affiliations:** Department of Microbiology, University of Innsbruck, 6020 Innsbruck, Austria

**Keywords:** SARS-CoV-2, storage, wastewater, stability, freezing

## Abstract

SARS-CoV-2 wastewater epidemiology suffers from uncertainties concerning sample storage. We show the effect of storage of wastewater on the detectable SARS-CoV-2 load. Storage at 4 °C up to 9 days had no significant effect, while storage at −20 °C led to a significant reduction in gene copy numbers.

**Highlights:**

- Raw wastewater samples can be stored up to 9 days at 4°C for SARS-CoV-2 wastewater surveillance
- Freezing of wastewater sample dramatically decreases RT-qPCR signal of SARS-CoV-2 from wastewater.

---

In the context of the global COVID19 pandemic, the quantification of severe acute respiratory syndrome coronavirus 2 (SARS-CoV-2) fragments in wastewater offers the opportunity to monitor the level of infection in large populations, independent of apparent symptoms (Medema et al., 2020; Wu et al., 2020b). With the growing numbers of SARS-CoV-2 wastewater studies, we need comprehensive knowledge on common storage procedures for raw wastewater to generate valid data from sewage surveillance. The aim of this investigation was to compare the effect of the most common storage temperatures of wastewater samples, +4°C and −20°C (Ahmed et al., 2020c; Michael-Kordatou et al., 2020)), on detectability of SARS-CoV-2 gene copy numbers.

Therefore, we analyzed 24 h composite samples of raw influent wastewater from the wastewater treatment plant (WWTP) Zirl, Tyrol, Austria (19.04.2020, 30’000 population equivalents (PE)) and WWTP Siggerwiesen, Salzburg, Austria (04.10.2020, 680’000 PE). The former samples were pasteurized prior to analysis due to uncertainties of the safety status of wastewater at this time, while the latter remained unpasteurized. Pasteurization of the wastewater involved an exposure of the samples to 60 °C for 1.5 h prior to sample processing (Darnell et al., 2004). The investigated storage conditions are summarized in Table 1.

**Table 1:** Experimental design

| WWTP | sampling date | Pasteurization | Storage at -18 °C | Storage at 4 °C |
| --- | --- | --- | --- | --- |
| Zirl, Tyrol | 19.04.2020 | yes | 2 days | 0, 1, 3, 7 days |
| Siggerwiesen, Salzburg | 04.10.2020 | no | 3 days | 0, 2, 7, 9 days |

For SARS-CoV-2 RNA extraction, we modified the protocol from (Wu et al., 2020b). In a first step, larger particles were removed to decrease the amount of non-viral RNA and PCR-inhibitors. For this purpose, 40 – 70 mL of wastewater were transferred to centrifugation tubes and centrifuged at 4’500 g for 30 min at 4°C. To precipitate viral fragments, the resulting supernatant was immediately transferred into a fresh tube containing 10 % w/v polyethylene glycol (PEG) 8000 (CarlRoth, Karlsruhe, Germany) and 2.25 % w/v NaCl. The Reax2™ head over-head shaker (Heidolph, Schwabach, Germany) was used until both additives were dissolved within a few minutes. Subsequently, the samples were centrifuged at 12’000 g for 99 min at 4 °C to obtain a pellet containing the viral fragments. The supernatant was removed in two steps. First, most of the supernatant was carefully decanted, then, after additional centrifugation at 12’000 g for 5 min, a pipette was used to remove the remaining fluid.

Following the precipitation of the viral fragments, pellets from the Zirl samples were resuspended with 800 µL TRIzol® (Invitrogen, USA) and TRIzol®-chloroform extraction was performed. For the Salzburg samples, we substituted hazardous TRIzol® with 800 µl lysis-buffer (Monarch™ total RNA Miniprep Kit, NewEnglandBiolabs, Ipswich, USA). The aqueous, pale phase from TRIzol®-chloroform extraction or the pellet resuspended in lysis-buffer were purified according to manufacturer protocol of the Monarch™ total RNA Miniprep Kit with non-enzymatic gDNA-removal. RNA was eluted in 40 µl RNase free water.

RNA concentrations of the templates were quantified via Nanodrop and extracts with RNA concentrations above 200 ng µL^−1^ were diluted as needed. RNA copy numbers were determined using the N1 primers/probe according the CDC-protocol (CDC, 2020) targeting the nucleocapsid-gene of SARS-CoV-2. RT-qPCR reactions contained per 20 µL: 10 µL Luna Universal Probe One-Step Reaction Mix (2X) from NEB, 1 µL Luna WarmStart^®^ RT Enzyme Mix (20X) from NEB, 0.8 µL primer (final concentration 0.4 µM), 0.4 µL probe (final concentration 0.2 µM), 2 µL PCR grade water, and 5 µL template. Analyses were conducted on a RotorGene cycler (Qiagen, Hilden, Germany). After an initial reverse transcription at 55 °C for 10 min, followed by 95°C for 1 min of denaturation, 45 cycles of 95°C for 10 sec and 60 °C for 40 sec were performed. To calculate copy numbers, a plasmid standard containing the N-gene of SARS-CoV-2 (2019-nCoV_N_Positive Control, IDT, Leuven, Belgium) was used. All variants were processed in parallels (n≥3) and were tested for significant difference against day 0 using the Mann-Whitney-U-Test (α = 0.05) in the software package Past 4.03 (Hammer et al., 2001). Within all samples stored at 4°C, variation coefficients spanned from 2% to 51% with a median of 37%. This variance inhomogeneity may be explained by the heterogeneity of the influent wastewater and by the accumulation of inaccuracies during the multi-step extraction protocol. Large variance and inhomogeneity were reported earlier by Wu et al (Wu et al., 2020a) and seem to be independent from the method of viral fragments concentration as reported by Ahmed et al (Ahmed et al., 2020b).

Short-time storage up to 9 days of wastewater at 4 °C had no significant effect on the number of detectable SARS-CoV-2 fragments (Figure 1 and Figure 2). These findings are in accordance with earlier studies on enveloped viruses (Casanova et al., 2009; Muirhead et al., 2020). In contrast to our storage-experiments, Ahmed et al. (2020a) choose a spike-in approach, using high loads of gamma irradiated SARS-CoV-2 virions (approximately 6.7×10^5^ gc ml^−1^) and stated a decay-rate of approximately 8% per day at 4°C.

**Figure 1:**
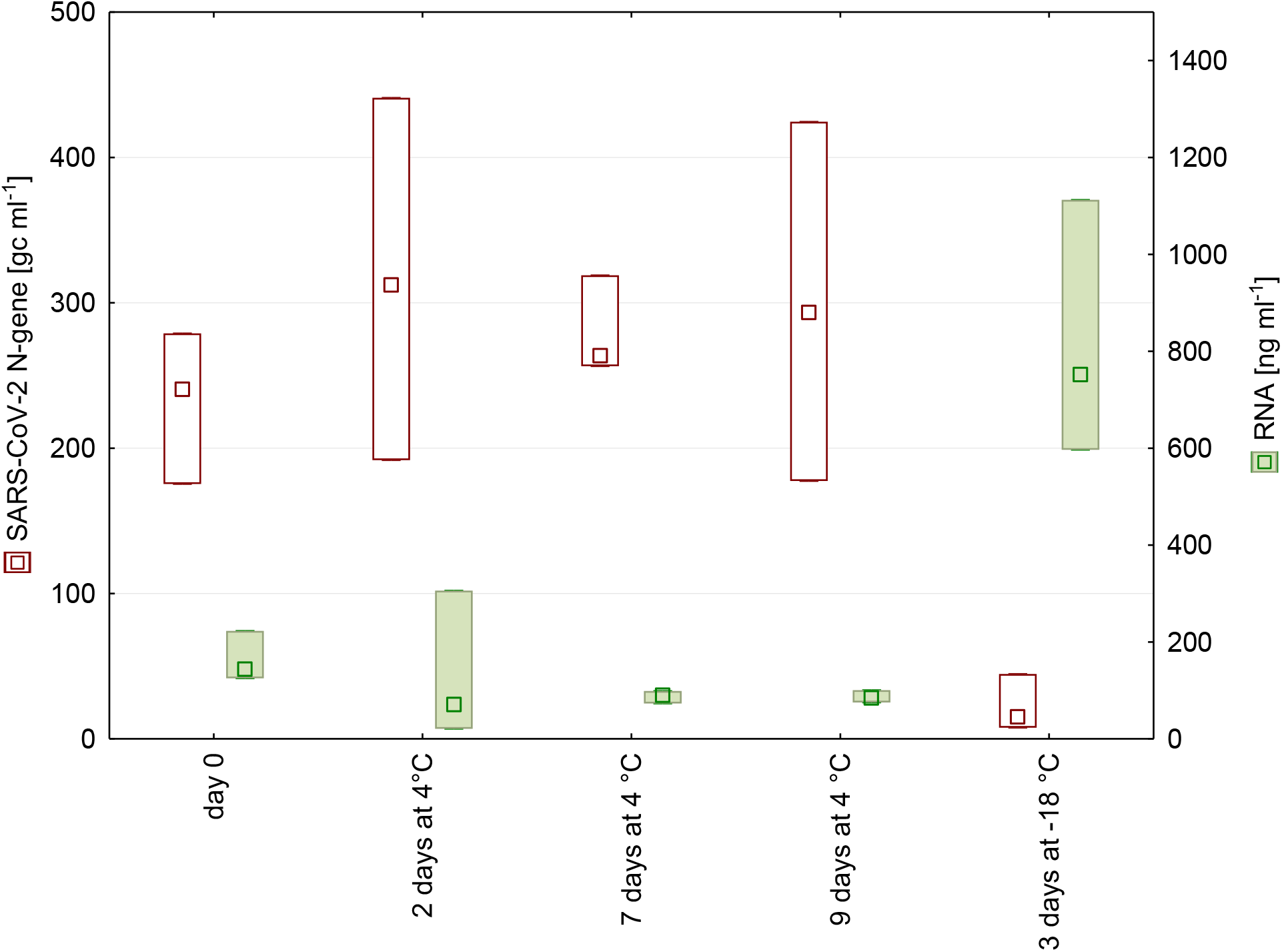
N-gene copy numbers and RNA concentration detected in wastewater from WWTP in Salzburg after 0, 2, 7, and 9 days of storage at 4 °C as well as after freezing. (n = 4, Median, Box: min-max).

**Figure 2:**
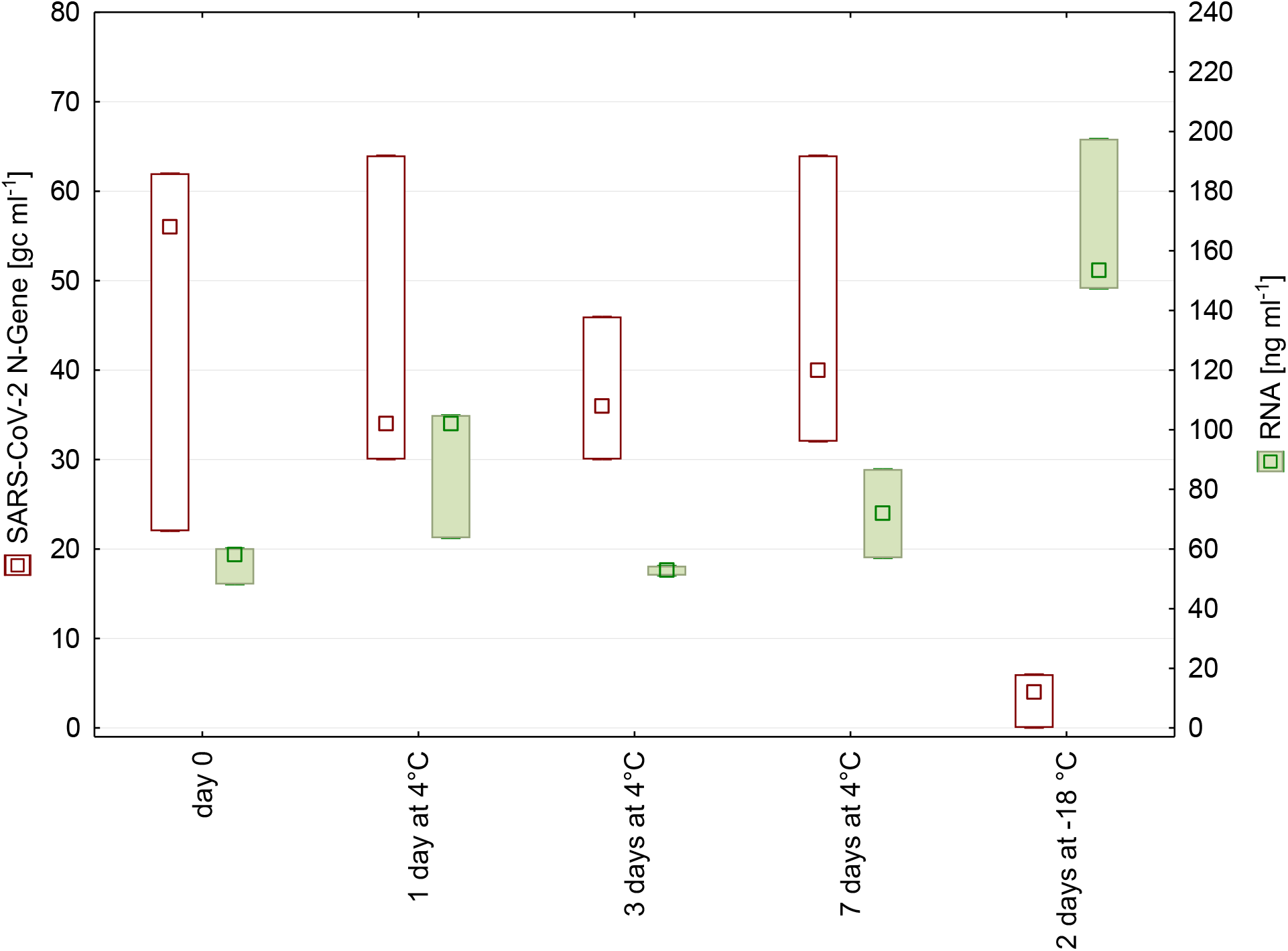
N-gene copy numbers and RNA concentration detected in wastewater from WWTP in Tyrol after 0, 1, 3, and 7 days of storage at 4 °C as well as after freezing. (n = 3, Median, Box: min-max).

Freezing/thawing of samples led to a significant loss of signal. A possible reason is that the freeze-thaw cycle disrupts cells, which is also reflected in increased RNA concentration in frozen samples (Figure 1 and Figure 2). The release of cell contents possibly includes also proteases and RNases, which may impair subsequent SARS-CoV-2 detection.

In conclusion, we recommend storing wastewater samples for SARS-CoV-2 analysis at 4 °C upon analysis and not freezing them.

## Data Availability

All data generated or analysed during this study are included in this published article.

## Acknowledgements

We would like to thank the WWTPs Zirl and Siggerwiesen very much for their collaboration and the supply with samples.

## Contributions

Rudolf Markt: Investigation, Methodology, Formal analysis, Visualization, Writing - original draft; Writing – review & editing;

Nina Lackner: Writing - original draft; Writing - review & editing; Data curation; Formal analysis Markus Mayr: Laboratory work, Project administration

Evelyn Peer: Laboratory work

Heribert Insam: conceptualization, manuscript editing

Andreas Wagner: Conceptualization, Writing - review & editing

